# Investigating Neural Dysfunction with Abnormal Protein Deposition in Alzheimer’s Disease Through Magnetic Resonance Spectroscopic Imaging, Plasma Biomarkers, and Positron Emission Tomography

**DOI:** 10.1101/2023.04.23.23288989

**Authors:** Kiwamu Matsuoka, Kosei Hirata, Naomi Kokubo, Takamasa Maeda, Kenji Tagai, Hironobu Endo, Keisuke Takahata, Hitoshi Shinotoh, Maiko Ono, Chie Seki, Harutsugu Tatebe, Kazunori Kawamura, Ming-Rong Zhang, Hitoshi Shimada, Takahiko Tokuda, Makoto Higuchi, Yuhei Takado

**Affiliations:** Department of Functional Brain Imaging, Institute for Quantum Medical Science, Quantum Life and Medical Science Directorate, National Institutes for Quantum Science and Technology, Chiba, Japan; Department of Psychiatry, Nara Medical University, Nara, Japan; QST Hospital, Quantum Life and Medical Science Directorate, National Institutes for Quantum Science and Technology, Chiba, Japan; Neurology Clinic, Chiba, Chiba, Japan; Institute for Quantum Life Science, Quantum Life and Medical Science Directorate, National Institutes for Quantum Science and Technology, Chiba, Japan; Department of Advanced Nuclear Medicine Sciences, Institute for Quantum Medical Science, Quantum Life and Medical Science Directorate, National Institutes for Quantum Science and Technology, Chiba, Japan; Center for integrated human brain science, Brain Research Institute, Niigata University, Niigata, Japan

**Keywords:** alzheimer’s disease, glutamate, magnetic resonance spectroscopy, positron emission tomography, neurofilament light chain plasma levels

## Abstract

In Alzheimer’s disease (AD), aggregated abnormal proteins induce neuronal dysfunction. Despite the evidence supporting the association between tau proteins and brain atrophy, further studies are needed to explore their link to neuronal dysfunction in the human brain. To clarify the relationship between neuronal dysfunction and abnormal proteins in AD-affected brains, we conducted magnetic resonance spectroscopic imaging (MRSI) and assessed the neurofilament light chain plasma levels (NfL). We evaluated tau and amyloid-β depositions using standardized uptake value ratios (SUVRs) of florzolotau (18F) for tau and ^11^C-PiB for amyloid-β positron emission tomography in the same patients. Heatmaps were generated to visualize Z scores of glutamate to creatine (Glu/Cr) and N-acetylaspartate to creatine (NAA/Cr) ratios using data from healthy controls. In AD brains, Z score maps revealed reduced Glu/Cr and NAA/Cr ratios in the gray matter, particularly in the right dorsolateral prefrontal cortex (rDLPFC) and posterior cingulate cortex (PCC). Glu/Cr ratios were negatively correlated with florzolotau (18F) SUVRs in the PCC, and plasma NfL levels were elevated and negatively correlated with Glu/Cr (P = 0.040, r = −0.50) and NAA/Cr ratios (P = 0.003, r = −0.68) in the rDLPFC. This suggests that the abnormal tau proteins in AD-affected brains play a role in diminishing glutamate levels. Furthermore, neuronal dysfunction markers including Glu/tCr and NAA/tCr could potentially indicate favorable clinical outcomes. Using MRSI provided spatial information about neural dysfunction in AD, enabling the identification of vulnerabilities in the rDLPFC and PCC within the AD’s pathological context.

## 1. Introduction

Alzheimer’s disease (AD) is a progressive neurodegenerative disease often characterized by initial memory impairment following a cognitive decline (Jack et al., 2013). A recent study reported the prevalence of clinical AD in the United States to be 11.3% and predicted further increases in the next four decades (Rajan et al., 2021). AD is a very burdensome disease, ranking fourth-highest in years of life lost in 2016 (Mokdad et al., 2018). The neuropathologic hallmark of AD is neuronal loss paralleled by the distribution of neurofibrillary tangles composed of filamentous tau proteins and amyloid plaques (DeTure and Dickson, 2019). The limited cognitive improvement observed upon removal of amyloid plaques with human monoclonal antibodies (van Dyck et al., 2023) suggests that therapeutic approaches that address other factors, including abnormal tau protein and neuronal dysfunction, may be necessary. Although the relationship between abnormal tau protein and brain atrophy has been established through previous human brain imaging studies (Shimada et al., 2017), understanding the manner in which neuronal dysfunction is related to abnormal proteins *in vivo* is crucial for the development of disease-modifying treatments rooted in the fundamental pathophysiology. To the best of our knowledge, only a small number of studies have investigated this relationship in cognitively unimpaired older adults (Kara et al., 2022).

Proton magnetic resonance spectroscopy (MRS) can potentially inform us about neuronal dysfunction in diseased brains using two major metabolites, N-acetylaspartate (NAA) and glutamate (Glu). While NAA has been employed as a primary neuronal marker of viability (Rae, 2014), Glu measured by MRS could be a marker of metabolic activity (Rae, 2014) in the brain due to its relationship with glucose metabolism (Sugiura et al., 2014). Studies have reported decreased Glu levels in the gray matter in patients with AD (reviewed in (Graff-Radford and Kantarci, 2013)). Furthermore, a recent meta-analysis of AD patients found reduced Glu levels in the posterior cingulate cortex (PCC), accompanied by reduced NAA levels (Liu et al., 2021). It is speculated that Glu may be an important metabolite in AD with a potential association with tau proteins; however, it has not been examined outside of the PCC using MRS, and the mechanism by which its relationship to abnormal proteins differs from NAA is not fully understood. This noninvasive modality can be combined with other modalities, such as positron emission tomography (PET) to evaluate brain conditions from multiple aspects.

PET imaging has enabled the evaluation of tau and amyloid-β deposition in AD pathology (Maschio and Ni, 2022). We reported that florzolotau (18F) (^18^F-florzolotau) could visualize tau depositions with high contrast in patients with AD (Tagai et al., 2021). A recent study showed that tau aggregations were associated with neuronal dysfunction evaluated by NAA and Glu levels in the PCC in cognitively unimpaired older adults (Kara et al., 2022). In the previous study, individuals with only slight abnormal protein accumulations were not excluded due to an absence cutoff of the PET standardized uptake value ratio (SUVR). Conversely, to our knowledge, no *in vivo* imaging studies investigated the associations of NAA and Glu levels with tau aggregations in patients who developed AD.

In this study, we utilized MRS imaging (MRSI) of the cingulate gyrus of patients with AD to test our hypothesis that patients with AD exhibited decreased levels of NAA and Glu, which are indicative of neuronal viability and metabolic activity, respectively, in the PCC and its relevant brain regions. In addition, we explored the connection between these MRSI measurements and the distribution of abnormal proteins measured by PET to elucidate their relationship. Moreover, we evaluated the clinical utility of MRS metabolites by examining the association between the blood levels of neurofilament light chain (NfL), which provides a sensitive measurement of neuroaxonal damage, and the MRS metabolites (Glu and NAA).

## 2. Material and methods

### 2.1. Participants

We recruited patients with AD, including mild cognitive impairment (MCI) due to AD and AD dementia from affiliated hospitals of the National Institute of Radiological Sciences (NIRS) and National Institutes for Quantum Science and Technology (QST) between April 2019 and September 2021. All the patients with MCI due to AD and AD dementia were diagnosed using the criteria for MCI defined by Petersen (Petersen et al., 1999) and the NINCDS-ADRDA Alzheimer’s Criteria as probable AD (McKhann et al., 1984), respectively. We also recruited age- and sex-matched healthy controls (HCs) from the volunteer association of the NIRS-QST. All the HCs had no history of neurologic or psychiatric disorders. At the screening, all the patients with AD and HCs were confirmed to be with and without amyloid deposition, respectively, by visual assessment of PET images of ^11^C-Pittsburgh compound-B (PiB) by three readers based on the standard method used in the Japanese Alzheimer’s Disease Neuroimaging Initiative (J-ADNI) study (Yamane et al., 2017). All the patients with AD also exhibited depositions of ^18^F-florzolotau in the neocortical and limbic cortices, aligning with our previous study using ^18^F-florzolotau PET in patients with AD (Tagai et al., 2021). We excluded one patient with AD and one HC because they did not complete the imaging protocol. Thirteen patients were exposed to cholinesterase inhibitors for AD (donepezil, n = 10; galantamine, n = 1; rivastigmine, n = 2). Meanwhile, we excluded one patient with AD who took memantine, a glutamate receptor antagonist. Finally, we enrolled 19 patients with AD and 27 HCs. We obtained written informed consent from all the participants and/or from spouses or other close family members when the patients were cognitively impaired. The QST Certified Review Board approved the current study.

All the participants were subjected to a series of standardized quantitative measurements of cognitive function (mini-mental state examination [MMSE] (Folstein et al., 1975) and Clinical Dementia Rating [CDR] (Morris, 1997) for AD patients) and neuropsychiatric symptoms (Apathy Scale (Starkstein et al., 1993) and Geriatric Depression Scale [GDS] (Lesher and Berryhill, 1994)). Additionally, all HCs and some AD patients completed the trail making test [TMT] (Tombaugh, 2004) (18 patients for TMT-A and 15 patients for TMT-B).

### 2.2. MRI / MRSI data acquisition and analysis

MRI and MRSI scans were performed using a 3.0-Tesla scanner (MAGNETOM Verio; Siemens Healthcare) equipped with a 32-channel receiving head coil. Three-dimensional T1-weighted images were acquired using a magnetization-prepared rapid gradient-echo sequence (repetition time (TR) = 2,300 ms; echo time (TE) = 1.95 ms; TI = 900 ms; field of view (FOV) = 250 mm; flip angle = 9°; acquisition matrix = 256 × 256; and axial slices of 1 mm thickness). We also performed a two-dimensional MRSI with point-resolved spectroscopy (PRESS) pulse sequence with the following parameters: TE = 30 ms, TR = 2000 ms, 3 averages, 16 × 16 chemical shift imaging matrix, and slice thickness = 15 mm. Every single volume of interest (VOI) was 10 × 10 × 15 mm^3^ (FOV = 160 × 160 mm^2^ including the measurement volume [mVol] = 80 × 80 × 15 mm^3^). We applied 3D Shim (Syngo MR version for VD13A, Siemens, Erlangen, Germany) and subsequently performed manual shimming to reduce the linewidth of the water spectrum in magnitude mode below 25 Hz. Outer-volume suppression (Tkác et al., 2001) and water suppression were enhanced through T1 effects (WET) (Ogg et al., 1994). A trained operator placed the multivoxel VOIs at the level of the cingulate gyrus (Figure 1).

**Fig. 1.**
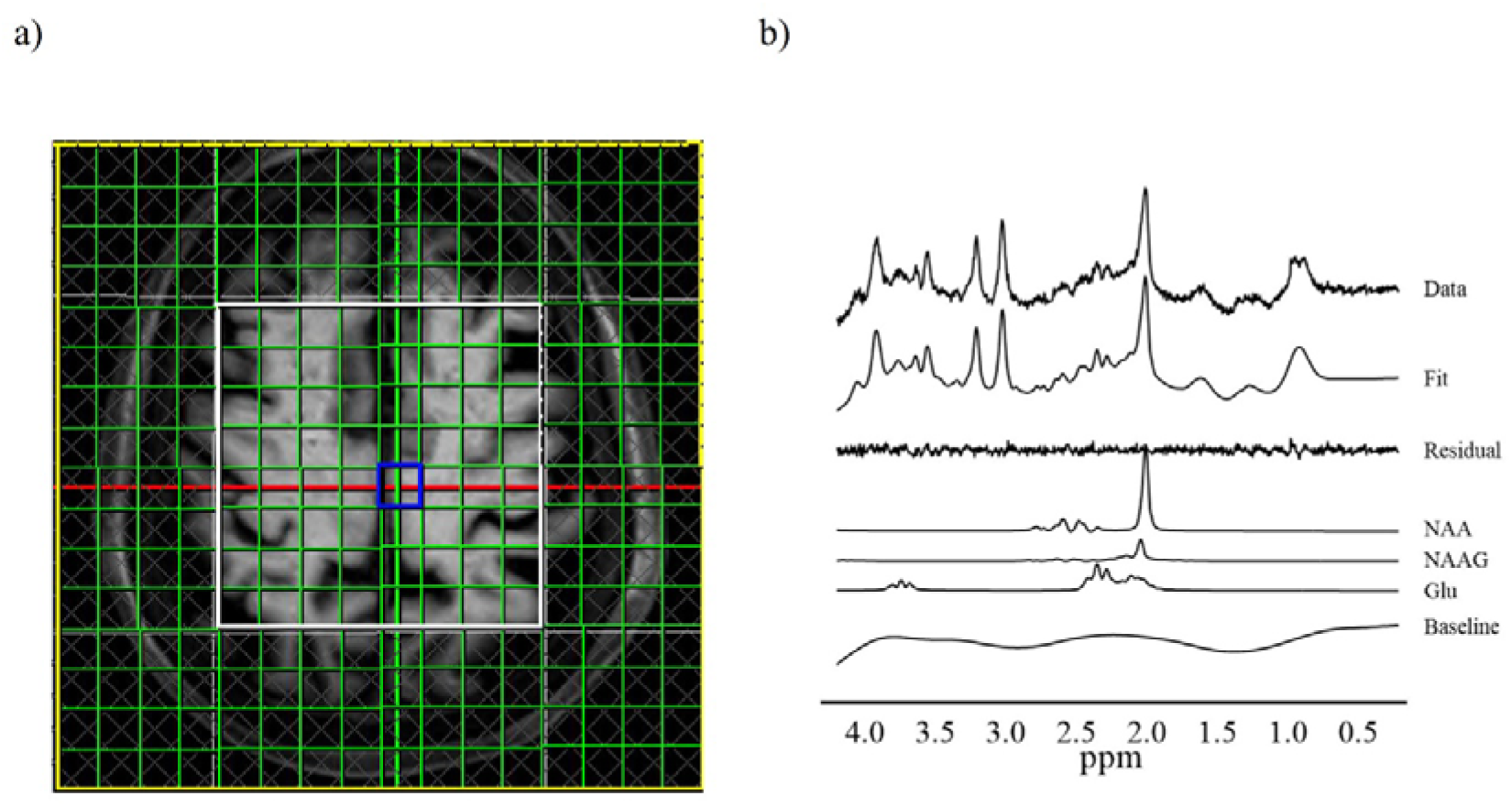
Multiple voxels of interest VOIs and representative spectrum of MRSI. a) We placed the multivoxel VOIs at the level of the cingulate gyrus. We obtained usable spectra from the 10 × 10 × 15 mm^3^ voxels (green squares) within the PRESS excitation volume (white box). b) An example of spectral data, spectral fit, fit residual, baseline, and individual metabolite fits obtained from the voxel (blue voxel). Glu, glutamate; NAA, N-acetylaspartate; NAA, N-acetylaspartate; NAAG, N–acetyl–aspartyl–glutamate; MRSI, magnetic resonance spectrum imaging; PRESS, point-resolved spectroscopy; VOI, volume of interest

### 2.3. PET procedures

All the participants were examined using ^11^C-PiB and ^18^F-florzolotau PET. ^11^C-PiB and ^18^F-florzolotau were radiosynthesized by the Department of Radiopharmaceuticals Development at NIRS following our previous studies’ protocols (Maruyama et al., 2013; Tagai et al., 2021). The ^18^F-florzolotau (injected dose: 185.7 ± 6.9 MBq, molar activity: 248.6 ± 76.3 GBq/μmol) PET scan was performed using Biograph mCT flow system (Siemens Healthcare). The images were reconstructed with filtered back projection. Meanwhile, the ^11^C-PiB (injected dose: 536.1 ± 61.1 MBq, molar activity: 87.2 ± 24.6 GBq/μmol) PET scan was performed using ECAT EXACT HR+ scanner (CTI PET Systems, Inc.), Biograph mCT flow system (Siemens Healthcare), or Discovery MI (GE Healthcare). All the PET scan images were reconstructed with filtered back projection.

### 2.4. MRSI data analysis

We analyzed MRSI data using LCModel software (Stephen Provencher, Inc) (Provencher, 1993) for a linear combination of model spectra provided in a basis set. We evaluated the referencing metabolite ratios of total NAA (tNAA; NAA + N–acetyl–aspartyl–glutamate [NAAG]) and Glu to total creatine (Cr) (tCr; Cr + PCr), because tCr has been widely used as an internal reference in human studies (Wilson et al., 2019). The signal-to-noise ratio (SNR) was obtained using an NAA peak height at 2.01 ppm divided by the standard deviation (SD) of noise. For all spectra, LCModel quantification was performed on a spectral window between 0.2 and 4.2 ppm. We excluded spectra data of the most dorsal 8 voxels, based on the quality control criteria (SNR < 5, full width at half maximum [FWHM] > 0.143 ppm) (Zhou et al., 2018). Additionally, data from one HC were excluded because of strong lipid contamination.

### 2.5. PET image processing

We applied motion correction and coregistration to the corresponding participant’s T1-weighted images for PET data using PMOD^®^ software ver. 3.8 (PMOD Technologies Ltd., Zurich, Switzerland). SUVR images were generated with cerebellar gray matter (GM) as a reference region, using ^18^F-florzolotau PET data at 90–110-min post-injection and ^11^C-PiB PET data at 50–70 min post-injection. For the reference region, we performed surface-based cortical reconstruction and volumetric subcortical segmentation of T1-weighted images with FreeSurfer software (version 6.0.0; http://surfer.nmr.harvard.edu). We defined regions of interest (ROIs) of the cerebellar cortex (Fischl et al., 2002) (excluding the vermis (Greve et al., 2016)) as described in our previous study (Matsuoka et al., 2022).

We defined target ROIs corresponding to the multivoxel VOIs of the measurement volume by MRSI. The multivoxel VOIs were composed 8 × 8 voxels of 80 × 80 × 15 mm^3^. We manually performed coregistration of the multivoxel VOIs to the corresponding participant’s T1-weighted images using screenshot images of the MRSI VOI placements as a reference (Figure S1). We evaluated ^18^F-florzolotau and ^11^C-PiB SUVRs in the multivoxel VOIs.

### 2.6. Immunoassay protocols

We measured plasma levels of the NfL from the blood samples of 26 HCs and 17 patients with AD. We applied an HD-X Simoa analyzer with reagents from a single lot using the Simoa NF-light assay, according to the protocol issued by the manufacturer (Quanterix, Lexington, MA, USA). All samples were analyzed in duplicate on one occasion.

### 2.7. Statistical analysis

Independent samples t-tests and χ^2^-tests were used to compare baseline demographics, clinical characteristics, metabolite levels, NfL levels, and ^18^F-florzolotau and ^11^C-PiB SUVRs. After performing the Shapiro–Wilk test to assess whether data followed normal distribution or not, Pearson correlation (for parametric data) or Spearman’s rank-order correlation (for nonparametric data) was used to evaluate the correlations between clinical characteristics, metabolite levels, NfL levels, and ^18^F-florzolotau and ^11^C-PiB SUVRs. All statistical tests were two-tailed, and P values of <0.05 indicated statistical significance. Statistical analyses were performed using IBM SPSS Statistics for Windows, version 25 (IBM Corp., Armonk, NY, USA).

We constructed heatmaps to visualize Z scores of ^18^F-florzolotau and ^11^C-PiB SUVRs and the metabolites to tCr ratios in patients with AD using data of HCs as a reference. In addition, we created a heatmap of Pearson correlation coefficients between the metabolites to tCr ratios and SUVRs of ^18^F-florzolotau and ^11^C-PiB SUVRs in patients with AD. We utilized GraphPad Prism version 8.4.3 (GraphPad Software) for heatmaps. Based on the results of the heatmaps, we chose VOI for further analysis to compare the metabolites with tCr ratios between patients with AD and HCs and the correlations with cognitive functions, NfL levels, and ^18^F-florzolotau and ^11^C-PiB SUVRs.

## 3. Results

### 3.1. Demographic and clinical profiles

The demographic and clinical profiles of patients with AD and HCs are summarized in Table 1. The study included eight patients with MCI and 11 with dementia. There were no statistically significant differences in age or sex between the two groups. The patients with AD showed worse cognitive functions in the total scores of MMSE and TMT-A and TMT-B than those of the HCs. There were no statistically significant differences in the symptoms of depression (GDS) or apathy (AS) between the two groups.

**Table 1.** Demographic characteristics.

|  | AD (n = 19) | HC (n = 26) |  |  |
| --- | --- | --- | --- | --- |
| | Mean (SD) or<br>frequency (%) | Mean (SD) or<br>frequency (%) | *t* or $\chi^2$ | P |
| Sex | | | $\chi^2 = 0.77$ | 0.28 |
| Female | 12 (63.2) | 13 (50) |  |  |
| Male | 7 (36.8) | 13 (50) |  |  |
| Age, years | 67.8 (9.4) | 67.8 (7.5) | *t* = -0.007 | 0.99 |
| Education, years | 14.1 (1.9) | 14.7 (1.5) | *t* = -1.15 | 0.26 |
| MMSE | 22.1 (4.0) | 28.8 (1.3) | *t* = -7.14 | <0.001 |
| TMT-A time <sup>a</sup> | 69.9 (31.5) | 35.5 (9.2) | *t* = 4.50 | <0.001 |
| TMT-B time <sup>b</sup> | 197.0 (93.6) | 85.4 (32.0) | *t* = 4.47 | <0.001 |
| GDS | 3.4 (2.2) | 2.9 (2.8) | *t* = 0.65 | 0.52 |
| Apathy Scale | 13.5 (5.2) | 10.9 (7.5) | *t* = 1.32 | 0.19 |
| CDR global score |  | N/A | N/A | N/A |
| 0.5 | 8 (42.1) |  |  |  |
| 1 | 9 (47.4) |  |  |  |
| 2 | 2 (10.5) |  |  |  |

Note: AD, Alzheimer's disease; CDR, clinical dementia rating; GDS, geriatric depression scale; HC, healthy control; MMSE, mini-mental state examination; SD, standard deviation; TMT, trail making test
<sup>a</sup>We presented available TMT-A data of 18 patients with AD/MCI and 26 HC participants.
<sup>b</sup>We presented available TMT-B data of 15 patients with AD/MCI and 26 HC participants.

### 3.2. Heatmaps of the Z score means of ^18^F-florzolotau and ^11^C-PiB SUVRs

All the patients with AD demonstrated typical depositions of ^18^F-florzolotau in the neocortical and limbic cortices, in line with our previous study (Tagai et al., 2021) (Figure 2a). They also showed ^11^C-PiB depositions in the cortices corresponding to the J-ADNI study criteria (Yamane et al., 2017) (Figure 2b). The heatmaps of the Z score means of ^18^F-florzolotau and ^11^C-PiB SUVRs were markedly increased in the neocortical and limbic cortices (Figure 2c, d).

**Fig. 2.**
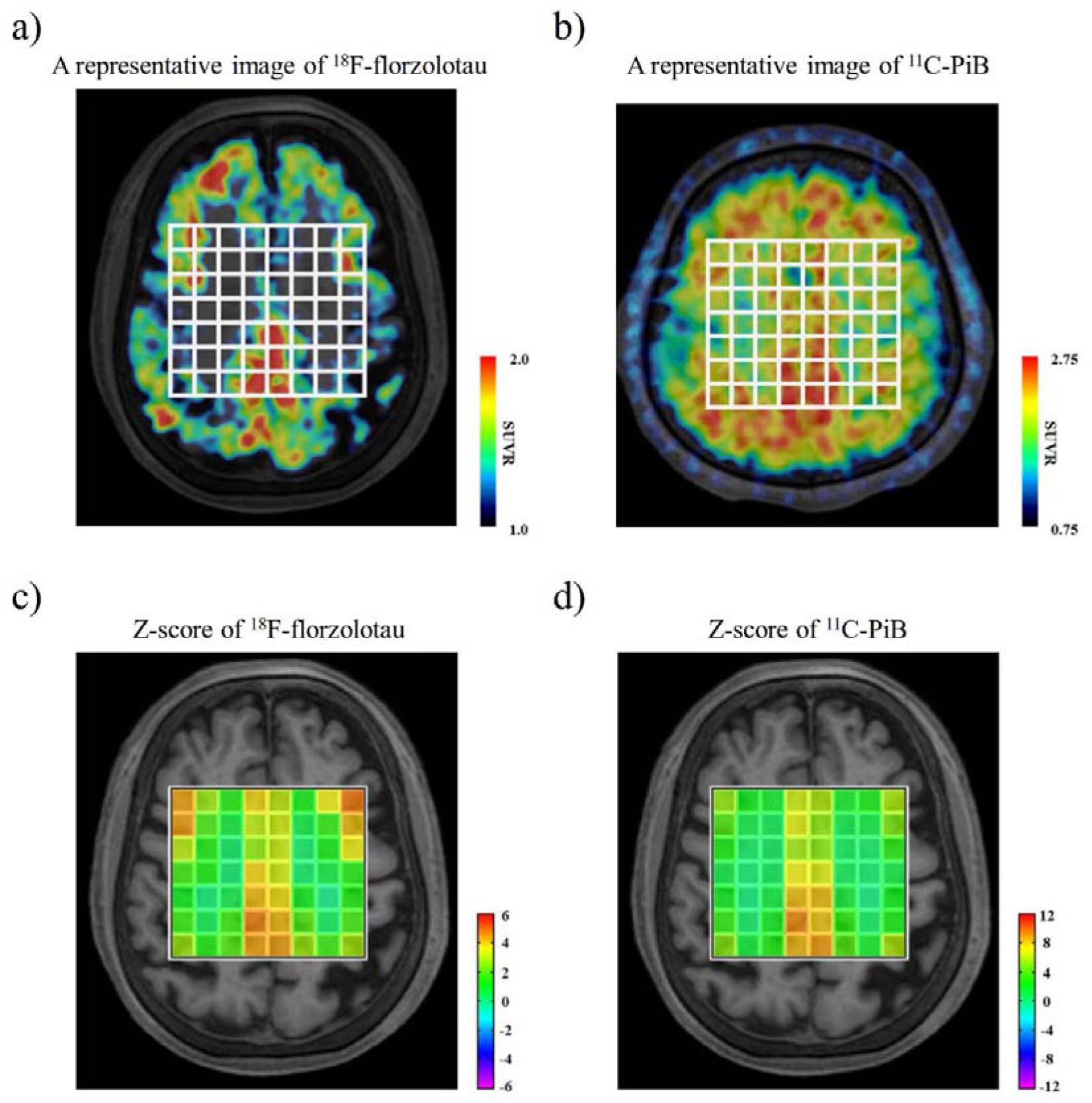
Representative PET SUVR images of a patient with AD and heatmaps of Z score means of ^18^F-florzolotau and ^11^C-PiB. a, b) A representative PET image of ^18^F-florzolotau (left) and ^11^C-PiB (right) in patients with AD showed accumulations in the neocortical and limbic cortices. c, d) Z score heatmaps of ^18^F-florzolotau (left) and ^11^C-PiB (right) in patients with AD also indicated marked increases in the neocortical and limbic cortices. AD, Alzheimer’s disease; PET, positron emission tomography; PiB, Pittsburgh compound-B; SUVR, standardized uptake value ratios

### 3.3. Heatmaps of Glu/tCr and tNAA/tCr ratios

The average Cramér–Rao lower bound of tNAA, Glu measurements, SNR, and FWHM in all voxels, except the most dorsal 8 voxels, were 2.7%, 8.2%, 26.0, and 0.048 ppm, respectively. The Cramér–Rao lower bounds of tNAA and Glu measurements were both <30% (Zhou et al., 2018). The heatmap showed decreased Z scores of Glu/tCr ratios in the GM, especially in the PCC and right dorsolateral prefrontal cortex (rDLPFC) (Figure 3a). The heatmap also demonstrated decreases in tNAA/tCr ratios in the GM, including the PCC and rDLPFC (Figure 3b).

**Fig. 3.**
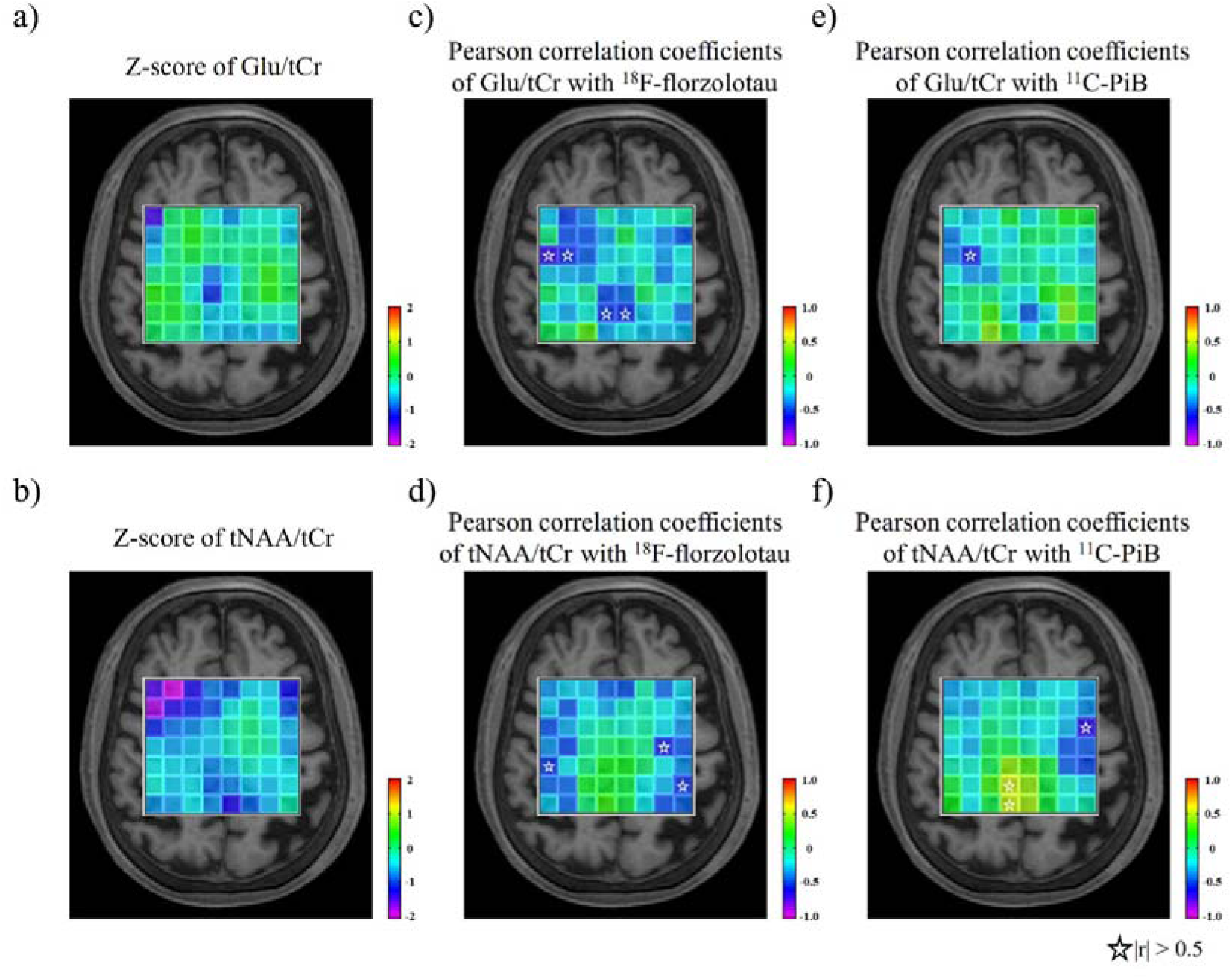
Heatmaps of Z score means of Glu/tCr and tNAA/tCr ratios and heatmaps of Pearson correlation coefficients with ^18^F-florzolotau and ^11^C-PiB SUVRs. a, b) The heatmap showed decreased Z scores of Glu (upper) and tNAA (lower) to tCr ratio in the GM, especially in the PCC and rDLPFC in patients with AD. c, d) The heatmaps of Pearson correlation coefficients displayed a marked negative correlation of Glu/tCr ratios with ^18^F-florzolotau SUVR in the PCC regions (upper), not but with ^11^C-PiB (lower) in patients with AD. e, f) The heatmaps of Pearson correlation coefficients indicated no extensive areas where ^11^C-PiB SUVRs were correlated with Glu (upper) and (lower) to the tCr ratio in patients with AD. Stars indicate that the absolute value of Pearson correlation coefficients is greater than 0.5. AD, Alzheimer’s disease; Cr, creatine; rDLPFC, right dorsolateral prefrontal cortex; Glu, glutamate; GM, gray matter; NAA, N-acetylaspartate; PCC, posterior cingulate cortex; PET, positron emission tomography; PiB, Pittsburgh compound-B; SUVR, standardized uptake value ratios.

The heatmaps of Pearson correlation coefficients of ^18^F-florzolotau and ^11^C-PiB SUVRs showed a marked negative correlation of Glu/tCr ratios with ^18^F-florzolotau SUVR in the PCC regions (Figure 3c–f).

### 3.4. Glu/tCr and tNAA/tCr ratios in the PCC and rDLPFC and their correlations with ^18^F-florzolotau and ^11^C-PiB SUVRs and cognitive batteries

Given that the heatmaps indicated decreased Glu/tCr and tNAA/tCr ratios in the PCC and rDLPFC, we subsequently analyzed the coalesced voxels covering the PCC and rDLPFC (Figure 4a), where their anatomical locations were confirmed using the atlas in FreeSurfer software (Figure S2). We found that Glu/tCr and tNAA/tCr ratios were both significantly decreased in the PCC and rDLPFC in patients with AD compared with that of the HCs (Glu/tCr ratios, P < 0.05 in the PCC and P < 0.005 in the rDLPFC; tNAA/tCr ratios, P < 0.05 in the PCC and P < 0.001 in the rDLPFC) (Figure 4b-e). We also found a correlation between tNAA/tCr ratios in the PCC and rDLPFC (r = 0.52, P = 0.023), but not with Glu/tCr (r = 0.16, P = 0.51).

**Fig. 4.**
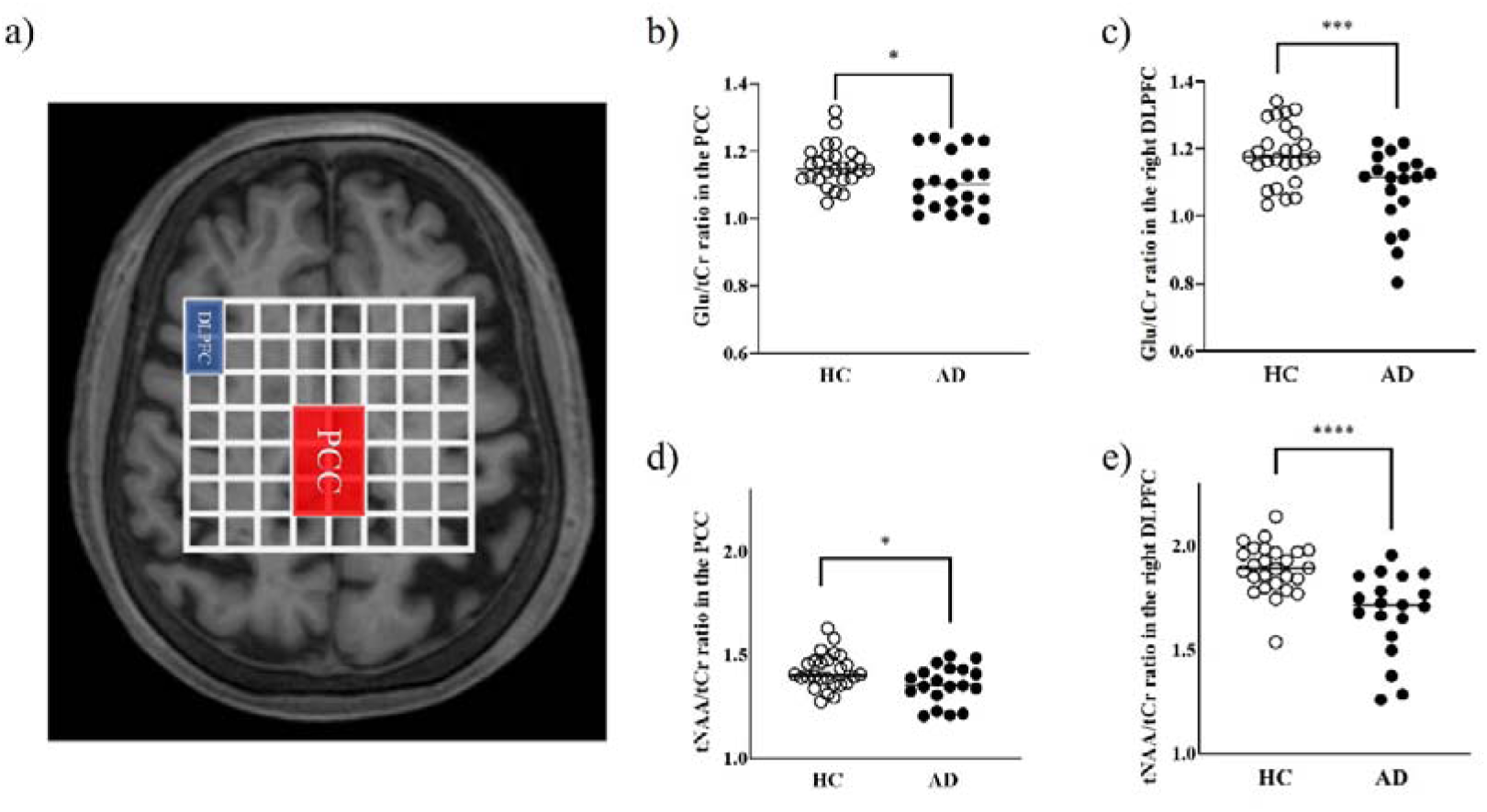
Representative image of combined voxels covering the PCC and rDLPFC, and scatterplots showing differences in the Glu/tCr and tNAA/tCr ratios between the patients with AD and HCs. a) Representative image of the combined voxels covering the PCC and rDLPFC (2 × 3 voxels of 20 × 30 × 15 mm^3^ for the PCC, and 1 × 2 voxels of 10 × 20 × 15 mm^3^ for the rDLPFC). b–e) Both the Glu/tCr and tNAA/tCr ratios were significantly decreased in the PCC and rDLPFC in patients with AD compared with those in the HCs (right upper, Glu/tCr ratios in the PCC; left upper, Glu/tCr ratios in the rDLPFC; right lower, tNAA/tCr ratios in the PCC; left lower, tNAA/tCr ratios in the rDLPFC). AD, Alzheimer’s disease; Cr, creatine; rDLPFC, right dorsolateral prefrontal cortex; Glu, glutamate; HC, healthy control; NAA, N-acetylaspartate; PCC, posterior cingulate cortex; PET, positron emission tomography; PiB, Pittsburgh compound-B. * P values are <0.05, *** P values are <0.005, **** P values are <0.001.

The heatmap revealed a negative correlation of ^18^F-florzolotau SUVRs with Glu/tCr ratios in the PCC but with not tNAA (Glu/tCr ratios, P = 0.046 and r = −0.46; tNAA/tCr ratios, P = 0.62 and r = 0.12) (Figure 5a, b). However, the correlation of ^18^F-florzolotau SUVRs with Glu/tCr ratios was not significant after Bonferroni correction for multiple comparisons, with P values set at <0.05/2 (where 2 is the number of ROIs). No significant correlations were observed with ^11^C-PiB SUVRs in the PCC (Glu/tCr ratios, P = 0.53 and r = −0.15; tNAA/tCr ratios, P = 0.11 and r = 0.38) (Figure 5c, d). This trend persisted after controlling for the scanners used for ^11^C-PiB PET (Glu/tCr ratios, P = 0.50 and r = −0.17; tNAA/tCr ratios, P = 0.14 and r = 0.36). Similarly, no significant correlations were observed with SUVRs of ^18^F-florzolotau and ^11^C-PiB in the rDLPFC (Glu/tCr ratios, P = 0.64 and r = −0.12; tNAA/tCr ratios, P = 0.19 and r = −0.32 for ^18^F-florzolotau; Glu/tCr ratios, P = 0.46 and r = −0.18; tNAA/tCr ratios, P = 0.17 and r = −0.33 for ^11^C-PiB) (Figure S3). These results remained nonsignificant after controlling for the scanners used for ^11^C-PiB PET (Glu/tCr ratios, P = 0.33 and r = −0.25; tNAA/tCr ratios, P = 0.093 and r = −0.41).

**Fig. 5.**
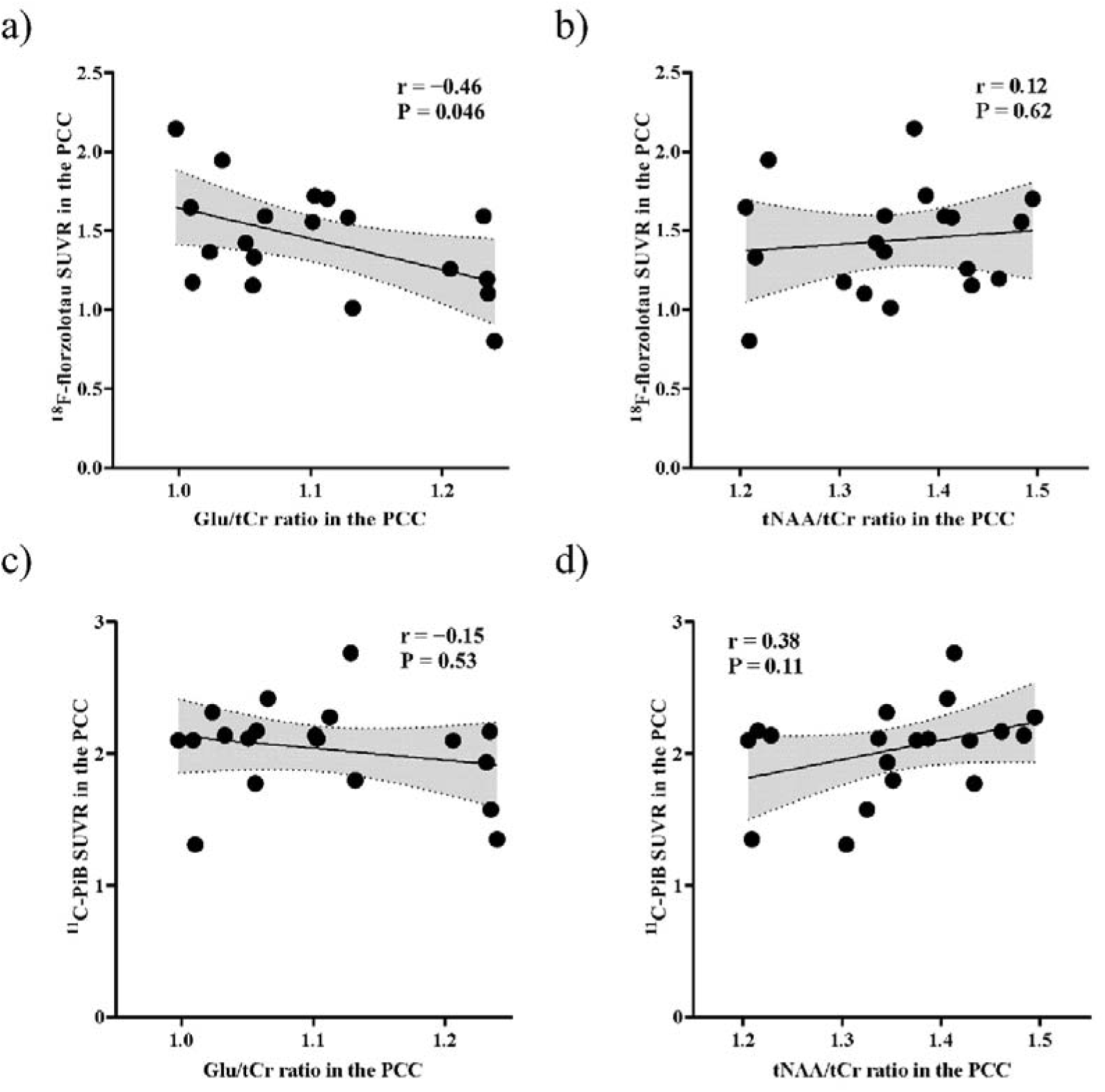
Scatterplot showing correlations of Glu/tCr and tNAA/tCr ratios with ^18^F-florzolotau and ^11^C-PiB SUVR in patients with AD. We found a negative correlation between ^18^F-florzolotau SUVRs and Glu/tCr ratios (upper left) in the PCC, but not with tNAA (upper right). There was no correlation of ^11^C-PiB SUVRs with Glu/tCr ratios (lower left) nor tNAA (lower right). AD, Alzheimer’s disease; Cr, creatine; Glu, glutamate; NAA, N-acetylaspartate; PCC, posterior cingulate cortex; SUVR, standardized uptake value ratios.

In the rDLPFC, Glu/tCr and tNAA/tCr ratios were positively correlated with MMSE total scores in patients with AD (Glu/tCr ratios, P < 0.001 and r = 0.72; tNAA/tCr ratios, P < 0.001 and r = 0.75) (Figure 6a, b), and these correlations remained significant after Bonferroni correction for multiple comparisons, with P values set at <0.05/2 (where 2 is the number of ROIs). The Glu/tCr and tNAA/tCr ratios in the PCC were not correlated with the MMSE total scores (Glu/tCr ratios, P = 0.15 and r = 0.35; tNAA/tCr ratios, P = 0.14 and r = 0.35) (Table S1). In patients with AD, we also found negative correlations between Glu/tCr ratios in the PCC and TMT-A time and between tNAA/tCr ratios in the rDLPFC and TMT-B time, and these correlations were also significant after Bonferroni correction for multiple comparisons, with P values set at <0.05/2 (where 2 is the number of ROIs) (Glu/tCr ratios, P = 0.002 and r = −0.68; tNAA/tCr ratios, P = 0.53 and r = −0.16 in the PCC; Glu/tCr ratios, P = 0.22 and r = −0.31; tNAA/tCr ratios, P = 0.050 and r = −0.47 in the rDLPFC for TMT-A; Glu/tCr ratios, P = 0.83 and r = −0.061; tNAA/tCr ratios, P = 0.55 and r = 0.17 in the PCC; Glu/tCr ratios, P = 0.95 and r = 0.017; tNAA/tCr ratios, P = 0.007 and r = −0.66 in the rDLPFC for TMT-B) (Figure 6c, d and Table S1).

**Fig. 6.**
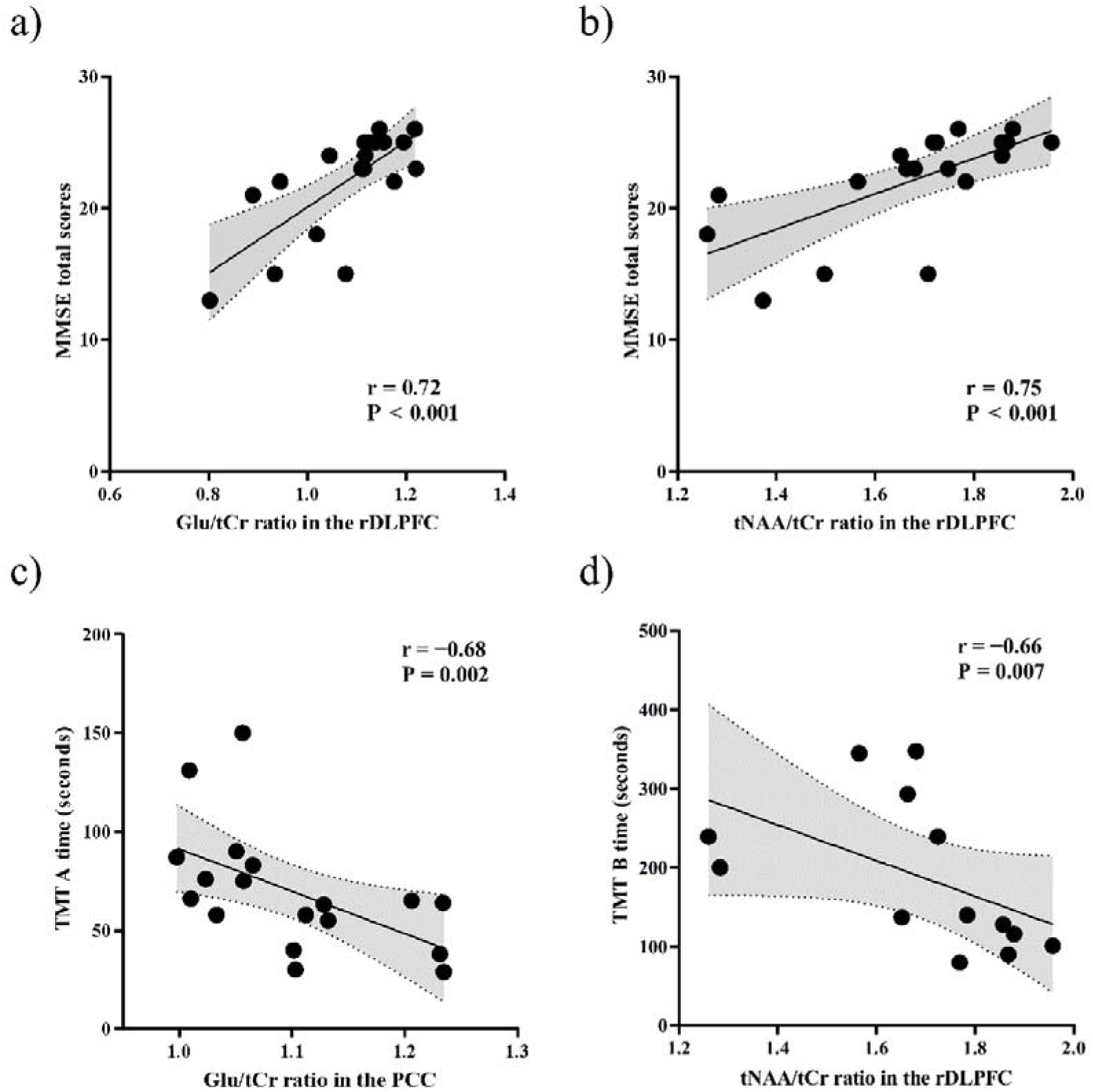
Scatterplot showing correlations of Glu/tCr and tNAA/tCr ratios with cognitive batteries in patients with AD. a, b) The Glu (upper left) and tNAA/tCr ratios (upper right) in the rDLPFC were positively correlated with MMSE total scores in patients with AD. c, d) We also found negative correlations between Glu/tCr ratios in the PCC and TMT-A time (lower left) and between tNAA/tCr ratios in the rDLPFC and TMT-B time (lower right) in patients with AD. AD, Alzheimer’s disease; Cr, creatine; rDLPFC, right dorsolateral prefrontal cortex; Glu, glutamate; NAA, N-acetylaspartate; PCC, posterior cingulate cortex

### 3.5. Correlations of Glu/tCr and tNAA/tCr ratios in the PCC and rDLPFC with blood NfL levels

We found higher NfL levels in the blood in patients with AD than that in the HCs (P < 0.005) (Figure 7a). We found negative correlations between blood NfL levels and Glu/tCr ratios (P = 0.040 and r = −0.50) and tNAA/tCr ratios (P = 0.003 and r = −0.68) in the rDLPFC. There were also trends of negative correlations between blood NfL levels and Glu/tCr ratios (P = 0.10 and r = −0.4 1) or tNAA/tCr ratios (P = 0.086 and r = −0.43) in the PCC, although these correlations did not reach a statistical significance level (Figure 7b–e). After Bonferroni correction for multiple comparisons, setting the P value threshold at <0.05/2 (where 2 is the number of ROIs), only the correlation between blood NfL levels and tNAA/tCr ratios in the rDLPFC remained significant.

**Fig. 7.**
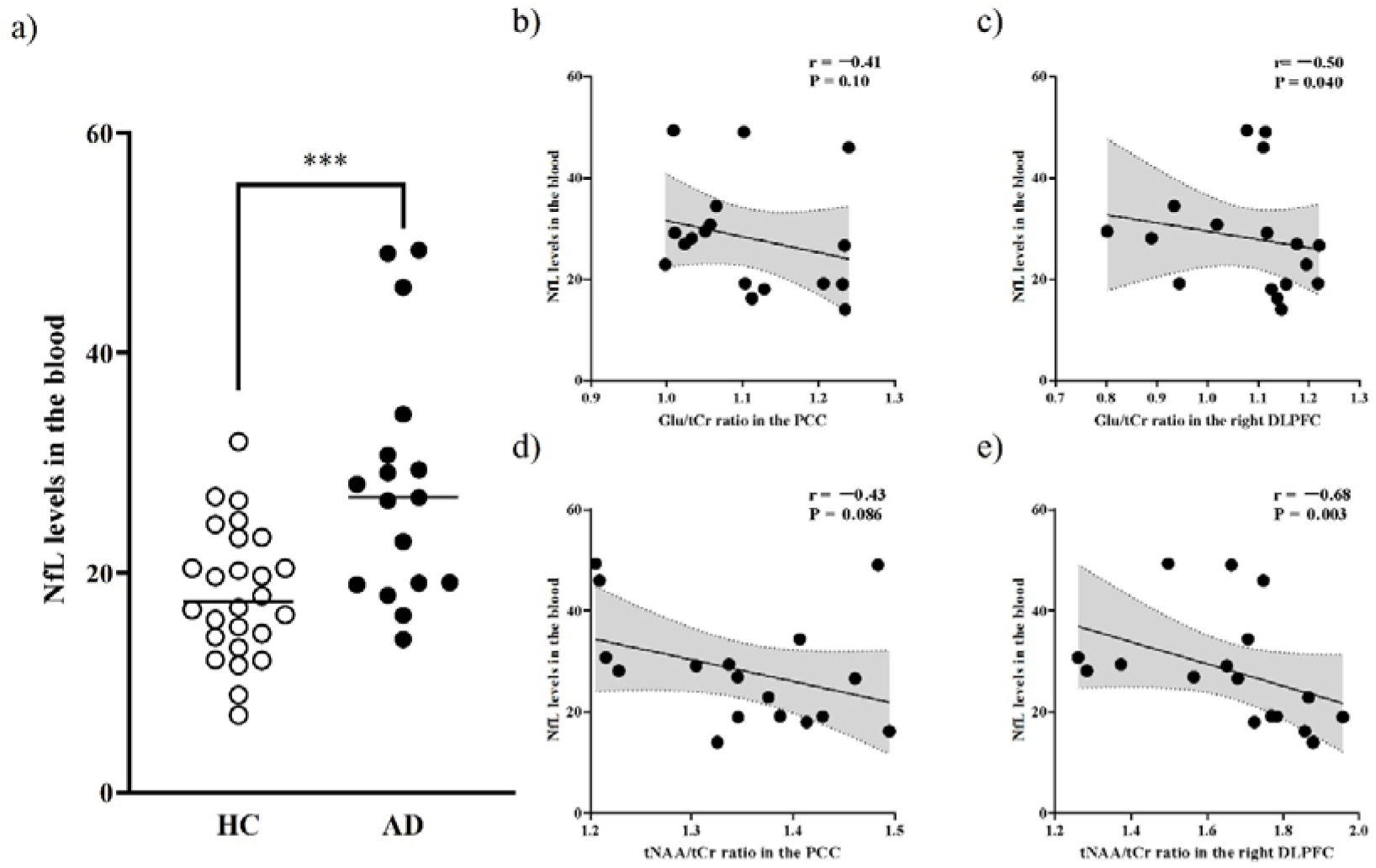
Scatterplots showing the comparison of blood NfL levels and the correlations of Glu/tCr and tNAA/tCr ratios with blood NfL levels in patients with AD. a) We found higher NfL levels in the blood of patients with AD than that in HCs. b–e) We found significant negative correlations between blood NfL levels and Glu/tCr ratios (upper right) and tNAA/tCr ratios (lower right) in the rDLPFC. There were also negative correlations between blood NfL levels and Glu/tCr ratios (upper left) and tNAA/tCr ratios (lower left) in the PCC, although these correlations did not reach a statistical significance level. AD, Alzheimer’s disease; Cr, creatine; rDLPFC, right dorsolateral prefrontal cortex; Glu, glutamate; NAA, N-acetylaspartate; NfL, neurofilament light chain; PCC, posterior cingulate cortex. *** P values are <0.005

## 4. Discussion

The MRSI technique revealed decreases in Glu/tCr and tNAA/tCr ratios in the PCC and rDLPFC, which were associated with impaired cognitive functions in patients with AD. Glu/tCr ratios, rather than tNAA/tCr ratios, exhibited a negative correlation with the levels of tau depositions in the PCC. Moreover, Glu/tCr and tNAA/tCr ratios in the rDLPFC were negatively associated with NfL levels, which provide a sensitive measurement of neuroaxonal damage.

The Glu/tCr and tNAA/tCr ratios in the PCC were lower in patients with AD compared with that of the HCs. While only the Glu/tCr ratio was found to correlate with ^18^F-florzolotau SUVRs in the PCC, the lack of correlation with the tNAA/tCr ratios requires further investigation. Glu is an essential metabolic biomarker, as previous research indicates that more than 50% of glucose eventually metabolizes to glutamate in the brain (Sugiura et al., 2014). Recent studies with meta-analysis have linked reduced brain glucose metabolism, mitochondrial dysfunction, and oxidative stress to pathology in AD, which induces synaptic damage and neuronal death (Ma et al., 2018; Song et al., 2021). In a previous study, the observed correlation between tau accumulation levels and Glu levels in the PCC of cognitively unimpaired patients is consistent with our findings (Kara et al., 2022). Tau pathology might reflect the alteration in neuronal functions compared with amyloid pathology, which reaches saturation in the early AD stage (Jack et al., 2013). Future preclinical studies could use a tauopathy mouse model to investigate the role of impaired brain glucose metabolism and mitochondrial function to elucidate the underlying mechanism of the relationship between decreased Glu levels and tau depositions. Furthermore, longitudinal studies in human and tau mouse models may clarify the time series of these associations.

In an exploratory MRSI study, reduced Glu/tCr and tNAA/tCr ratios were observed not only in the PCC but also in the rDLPFC, indicating the potential for the latter region to be a useful assessment region for MRS (Gozdas et al., 2022; Tumati et al., 2018). Prior studies on Glu levels in the DLPFC of patients with AD have reported inconsistent results, potentially due to the evaluation of Glx (Glu + glutamine [Gln]) rather than Glu alone. However, our measurement of Glu levels independent from Gln levels revealed reduced Glu levels in the rDLPFC of patients with dementia due to MCI and AD. Glu level differences between AD and HC were observed in the rDLPFC but not in the left dominant hemisphere (Glu/tCr ratios, P = 0.076 in the left DLPFC), although tNAA levels were significantly decreased in patients with AD compared with HCs (tNAA/tCr ratios, P < 0.001 in the left DLPFC). Although this study only captured changes in Glu/tCr ratios in the rDLPFC, the differences between the left and rDLPFC in patients with AD remain unclear. The results of a previous study utilizing ^18^F-FDG PET were in agreement with our investigations, indicating a decrease in metabolism of the DLPFC in the early stages of AD (Strom et al., 2022), particularly in the right hemisphere (Riederer et al., 2018). Nevertheless, MRSI is not without limitations, as chemical shift displacement may introduce ambiguity when interpreting interhemispheric discrepancies, especially considering the voxel on the rDLPFC at the edge of the MRSI volume. A previous study using a fat–water phantom reported a higher bias in chemical shift displacement error (CSDE) for the PRESS sequence compared with the sLASER sequence (28% vs. 10%, respectively) (Sourdon et al., 2021). An expert consensus paper on MRS also emphasized the limitation of CSDE in conventional sequences, such as the PRESS sequence, and indicated that this limitation is overcome by the sLASER sequence (Öz et al., 2020). Therefore, validation of the left–right difference of DLPFC function in AD should be further pursued using advanced MRS techniques, such as sLASER (Pradhan et al., 2015), which exhibits lower chemical shift displacement.

Conversely, our findings in the rDLPFC revealed positive associations between the Glu/tCr and tNAA/tCr ratios and the overall MMSE scores, while the tNAA/tCr ratio demonstrated an inverse correlation with TMT-B completion time. These results indicate that the outcomes in the rDLPFC are not a happenstance, but rather hold potential as a valuable appraisal domain for MRS, akin to the PCC. Given that the DLPFC has been shown to play a critical role in visual working memory (Li et al., 2017; Owen et al., 2005), it is plausible that reduced metabolic activity and neuronal survival in patients with AD may be linked to compromised working memory function. Our findings indicated that Glu/tCr and tNAA/tCr ratios may serve as potential clinical imaging markers for cognitive functions in patients with MCI due to AD.

Both MRSI and blood biomarkers showed neural dysfunctions in patients with AD, and as an alternative to PET, blood biomarkers and MRSI measurements are less invasive and less costly, making them potentially appropriate for use in primary care. Blood biomarkers, in particular, require less testing time. NfL has been established as a biomarker of neural damage in various neurological diseases, including AD, based on accumulating evidence (Khalil et al., 2018). MRSI is a noninvasive technique that provides spatial information on metabolite levels in the brain without requiring additional imaging modalities. However, the practical application of MRSI may be limited by the technical expertise required for imaging and data analysis. In this study, a single-slice MRSI approach was used to cover a portion of the brain region of interest, but recent technological advancements have enabled 3D imaging. Although resting-state functional MRI has been extensively utilized to assess the functional network of brain diseases, MRSI can directly evaluate neural viability and metabolic activity, and thus may provide useful complementary information for future brain network assessment from a different perspective.

The present study had limitations. We acquired ^11^C-PiB PET data using three scanners, and our ^11^C-PiB PET results should be interpreted with caution while considering the effects of different scanners (Albano et al., 2021; Dondi et al., 2022). Second, some correlations did not survive after correction for multiple comparisons, likely due to the small patient cohort. Third, at the time of the study, 10 patients with AD were taking donepezil, a medication reported to increase Glu/tCr ratios (Westman et al., 2009). Further studies with large sample sizes and adjustment for medications are needed to validate our findings. Fourth, we did not evaluate the fractions of GM, white matter, and CSF within the MRSI voxels in the present study. Further analysis on fraction control in the voxels is needed to draw more definitive conclusions.

## 5. Conclusions

The MRSI technique revealed neural dysfunctions, including metabolic activity, linked with tau aggregations in the PCC. MRSI has the potential to provide spatial information regarding neural functions, leading to our findings of deteriorated neural viability and metabolic activity in the rDLPFC of patients with AD. MRSI could qualify metabolic levels in several brain regions simultaneously. The robustness of MRSI data was demonstrated by its relationship with NfL, which is commonly used in blood biomarker studies. Together with resting-state functional MRI, these MRSI measurements may have the potential to elucidate brain network impairments.

## Author contributions

**Kiwamu Matsuoka:** Conceptualization, Data curation, Formal analysis, Funding acquisition, Investigation, Project administration, Visualization, Writing – original draft, Writing – review & editing. **Kosei Hirata:** Data curation, Formal analysis, Investigation, Resources, Validation, Writing – review & editing. **Naomi Kokubo:** Data curation, Investigation, Writing – review & editing. **Takamasa Maeda:** Data curation, Formal analysis, Investigation, Methodology, Writing – review & editing. **Kenji Tagai:** Data curation, Formal analysis, Investigation, Resources, Writing – review & editing. **Hironobu Endo:** Data curation, Formal analysis, Investigation, Writing – review & editing. **Keisuke Takahata:** Data curation, Investigation, Resources, Writing – review & editing. **Hitoshi Shinotoh:** Data curation, Investigation, Resources, Writing – review & editing. **Maiko Ono:** Methodology, Validation, Writing – review & editing. **Chie Seki:** Formal analysis, Methodology, Software, Writing – review & editing. **Harutsugu Tatebe:** Formal analysis, Methodology, Writing – review & editing. **Kazunori Kawamura:** Data curation, Investigation, Writing – review & editing. **Ming-Rong Zhang:** Supervision, Writing – review & editing. **Hitoshi Shimada:** Data curation, Investigation, Resources, Writing – review & editing. **Takahiko Tokuda:** Formal analysis, Funding acquisition, Methodology, Supervision, Writing – review & editing. **Makoto Higuchi:** Funding acquisition, Supervision, Writing – review & editing. **Yuhei Takado:** Conceptualization, Formal analysis, Funding acquisition, Project administration, Resources, Validation, Writing – original draft, Writing – review & editing.

## Data availability

The datasets generated during and/or analyzed during the current study are available from the corresponding author on reasonable request.

## Acknowledgments

The authors thank all patients and their caregivers for participation in this study, clinical research coordinators, PET and MRI operators, animal care technicians, radiochemists, and research ethics advisers at QST for their assistance in the current projects. We thank APRINOIA Therapeutics for kindly sharing the precursor of ^18^F-florzolotau. The authors acknowledge support for the recruitment of patients by Dr. Shigeki Hirano at the Chiba University and Dr. Yasumasa Yoshiyama, at Inage Neurology and Memory Clinic.

## Appendix A. upplementary data

Supplementary data to this article can be found online at ###.

## Ethics approval

This study was approved by the Institutional Review Board of the NIRS. Written informed consent was obtained from all patients and from their spouses or other close family members. The study was registered with the UMIN clinical Trials Registry (UMIN-CTR; number 000030248).

