## Supplemental_materials for "Investigating Neural Dysfunction with Abnormal Protein Deposition in Alzheimer’s Disease Through Magnetic Resonance Spectroscopic Imaging, Plasma Biomarkers, and Positron Emission Tomography"

### **Supplementary Materials**

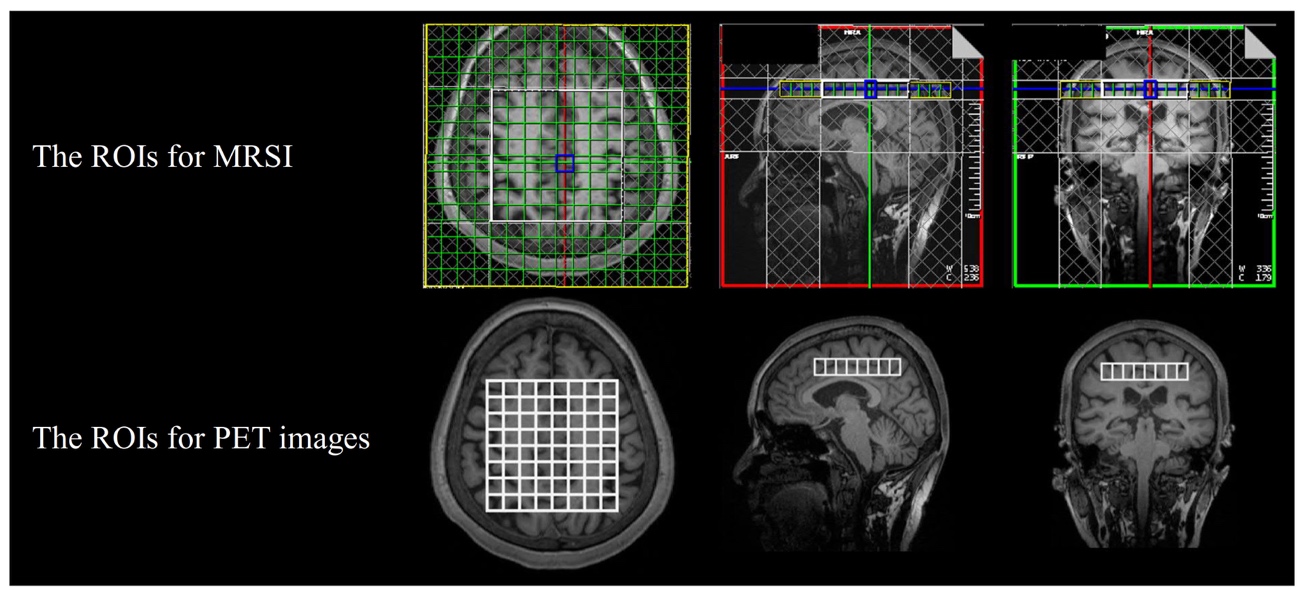

### **Figure S1.** Representative images of multiple VOIs for MRSI and PET images.

We manually performed coregistration of the multivoxel VOIs to the corresponding participant’s T1-weighted images, using the screenshot images of the MRSI VOIs placements as a reference.

MRSI, magnetic resonance spectrum imaging; PET, positron emission tomography; VOI, volume of interest

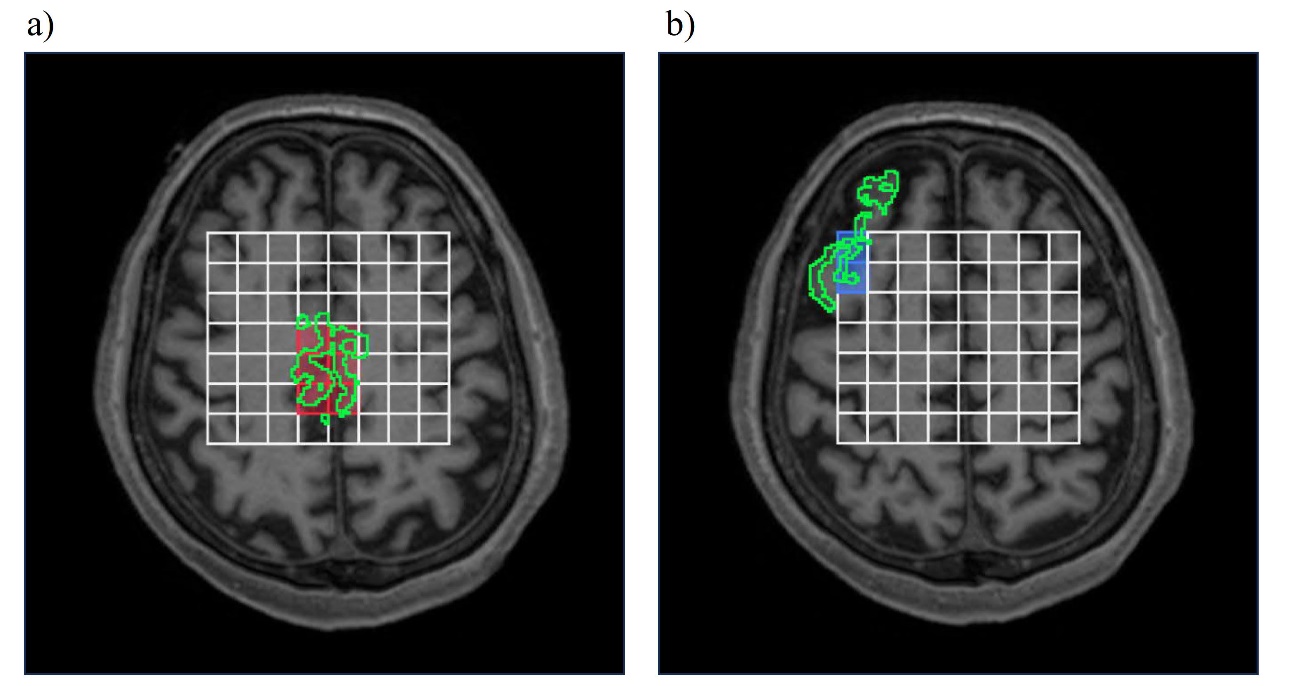

### **Figure S2**. Representative image of combined voxels covering the PCC and rDLPFC and their anatomical locations in the atlas of FreeSurfer software.

### Representative image of the combined voxels covering the PCC (red voxels) and rDLPFC (blue voxels). Anatomical locations were confirmed using the atlas in FreeSurfer software. ROIs (green areas) of (a) PCC and (b) rDLPFC were defined using atlases (Fischl et al., 2004; Klein and Tourville, 2012).

### rDLPFC, right dorsolateral prefrontal cortex; PCC, posterior cingulate cortex; ROI, regions of interest

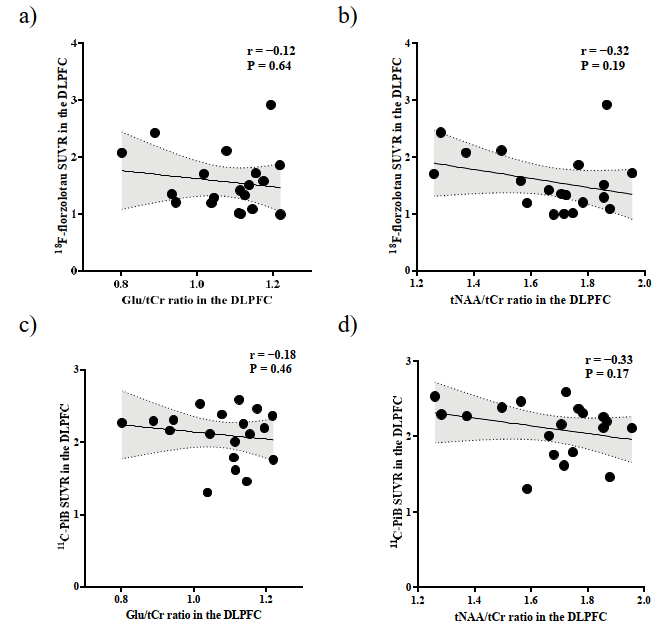

**Figure S3**. Scatterplot showing correlations of Glu/tCr and tNAA/tCr ratios with ^18^F-florzolotau and ^11^C-PiB SUVR in patients with AD in the right DLPFC.

No correlations were observed between ^18^F-florzolotau SUVRs and Glu/tCr ratios (upper left) or tNAA (upper right). ^11^C-PiB SUVRs were not correlated with Glu/tCr ratios (lower left) nor tNAA (lower right). AD, Alzheimer’s disease; Cr, creatine; Glu, glutamate; NAA, N-acetylaspartate; rDLPFC, right dorsolateral prefrontal cortex; SUVR, standardized uptake value ratios

### **Table S1. Correlations of the cognitive batteries with the Glu/Cr and NAA/Cr ratios in patients with AD**

|  | r (P) | | | |
| --- | --- | --- | --- | --- |
|  | PCC | | Right DLPFC | |
|  | Glu/tCr | tNAA/tCr | Glu/tCr | tNAA/tCr |
| MMSE total scores | 0.35 (0.15) | 0.35 (0.14) | 0.72 (<0.001) | 0.75 (<0.001) |
| TMT-A time | −0.68 (0.002) | −0.16 (0.53) | −0.31 (0.22) | −0.47 (0.050) |
| TMT-B time | −0.061 (0.83) | 0.17 (0.55) | 0.017 (0.95) | −0.66 (0.007) |

AD, Alzheimer’s disease; DLPFC, dorsolateral prefrontal cortex; Glu, glutamate; MMSE, mini mental state examination; NAA, N-acetylaspartate; PCC, posterior cingulate cortex; TMT, trail making test
